# The time-series ages distribution of the reported COVID-2019 infected people suggests the undetected local spreading of COVID-2019 in Hubei and Guangdong provinces before 19^th^ Jan 2020

**DOI:** 10.1101/2020.03.28.20040204

**Authors:** Chao Wu, Min Zheng

## Abstract

COVID-2019 is broken out in China. It becomes a severe public health disaster in one month. Find the period in which the spreading of COVID-2019 was overlooked, and understand the epidemiological characteristics of COVID-2019 in the period will provide valuable information for the countries facing the threats of COVID-2019. The most extensive epidemiological analysis of COVID-2019 shows that older people have lower infection rates compared to middle-aged persons. Common sense is that older people prefer to report their illness and get treatment from the hospital compared to middle-aged persons. We propose a hypothesis that when the spreading of COVID-2019 was overlooked, we will find more older cases than the middle-aged cases.

At first, we tested the hypothesis with 4597 COVID-2019 infected samples reported from 26^th^ Nov 2019 to 17^th^ Feb 2020 across the mainland of China. We found that 19^th^ Jan 2020 is a critical time point. Few samples were reported before that day, and most of them were older ones. Then samples were explosively increased after that day, and many of them were middle-aged people. We have demonstrated the hypothesis to this step.

Then, we grouped samples by their residences(provinces). We found that, in the provinces of Hubei and Guangdong, the ages of samples reported before 19^th^ Jan 2020 are significantly higher than the ages of samples reported after that day. It suggests the COVID-2019 may be spreading in Hubei and Guangdong provinces before 19^th^ Jan 2020 while people were unconscious of it.

At last, we proposed that the ages distribution of each-day-reported samples could serve as a warning indicator of whether all potential COVID-2019 infected people are found.

We think the power of our analysis is limit because 1. the work is data-driven, and 2. only ∼5% of the COVID-2019 infected people in China are included in the study. However, we believe it still shows some value for its ability to estimate the possible unfound COVID-2019 infected people.

## Introduction

COVID-2019 is broken out in China. It becomes a severe public health disaster in one month. 78630 is confirmed to be COVID-2019 infected people to the date of 27^th^ Feb 2020 in China^1^. With a considerable effort and abundant cost, China has controlled the disaster in the provinces out of Hubei. However, in the world, the number of COVID-2019 infected people is reaching 1000 in Korea and Japan, and it is entering or over 100 in Singapore and Italy until 26^th^ Feb 2020. The Chinese government and the Korean government have taken the strictest action to fight against COVID-2019 right now. A serious problem that the other governments facing the threats of COVID-2019 have to answer is: are all COVID-2019 infected people found? Dr. Tedros Adhanom Ghebreyesus has warned “we may only be seeing the tip of the iceberg” and that all countries “must use the window of opportunity” to contain the spread of the virus.

The older people have lower COVID-2109 infection rates. China CDC (Chinese Center for Disease Control and Prevention) has conducted the most extensive epidemiological analysis of COVID-2019 based on 72314 infected cases^3^. They found that 77.8% of infected cases are between 30 and 69; the cases above 70 or below 30 have lower infection rates. In our previous work (submitted to China CDC Weekly; Manuscript ID CCDCW200015), we found that the cases above 65 or below 20 tend to have lower COVID-2019 infection rates compared to the cases between 20 and 65, which is consistent with the epidemiological analysis of COVID-2019 conducted by China CDC.

Current research shows that older people tend to get severe clinical symptoms. The 80% of death cases of COVID-2019 are people with ages above 60^3^. Common sense is that the older people prefer to report their illness and get treatment from the hospital compared to the middle-aged persons and youths. We propose a hypothesis that when the spreading of COVID-2019 is overlooked, more older cases will be reported. Once they realize that COVID-2019 is spreading, people of different ages will pay more attention to their health and visit the doctors even they have few or no clinical symptoms.

We tested the hypothesis with 4597 infected samples reported from 26^th^ Nov 2019 to 17^th^ Feb 2020 across the mainland of China. We found the ages of the samples reported before 19^th^ Jan 2020 are significantly higher than the ages of samples reported after 19^th^ Jan 2020. We further group samples by their residences (provinces). We found that the ages of the samples reported before 19^th^ Jan 2020 are significantly higher than the ages of samples reported after that day in the provinces of Hubei and Guangdong. It suggests the COVID-2019 may be spreading in Hubei and Guangdong provinces before 19^th^ Jan 2020 while people were unconscious of it.

We further compared the ages of samples reported before 19^th^ Jan 2020 and the ages of samples reported each day after 19^th^ Jan 2020. We found that the ages of samples reported before 19^th^ Jan 2020 tend to be higher than the ages of the each-day-reported samples. We infer the ages of each-day-reported infected people could serve as a warning indicator of whether all potential COVID-2019 infected cases are found.

The work is data-driven. The power of the work is limit because only ∼5% of the COVID-2019 infected people in China are included in the study. However, we believe it still shows some value for its ability to estimate the possible un-found COVID-2019 infected people.

## Methods

“Liberation Daily · Shangguan news” has collected the cases of COVID-2019 infected people in China^2^. It recorded the ages, residences, and the dates on which they visited the doctors for four thousand five hundred ninety-seven infected cases in China to the time of 18:00 (Beijing time), 18^th^ Feb 2020. We modified his age as “50” for the person whose age is recorded as “50+”. We recorded their ages by year for the fifteen infants whose ages were marked by month. We selected the earlier date for the seven infected cases marked with multiple times on which they visited the doctors. Then we got four thousand five hundred ninety-seven affected cases with information about the ages, residences, and the dates on which they visited the doctors. We took them as samples and conducted analyses on them.

## Result

### The infection rate of samples between 30 and 60 is twice compared to the people above 60

We grouped samples by their ages and calculated the infection rates of samples in different groups. For each group, the infection rate is calculated as the number of samples in the group dividing the number of total samples (Figure 1A). We found the ones on the groups of 30-35, 35-40,40-45,45-50, 50-55, 55-60 have higher infection rates than the ones on the groups of 60-65,65-70,70-75,75-80, 80-85, 85-90, 90-95 and 95-100. The infection rate of samples between 30 and 60 (0.62) is twice more than the samples above 60 (0.19). In the epidemiological analysis conducted by China CDC, the percent of samples in the ages between 30 and 60 (0.59) is near twice the percent of samples in the ages above 60 (0.31). The older people have lower COVID-2109 infection rates.

**Figure 1.**
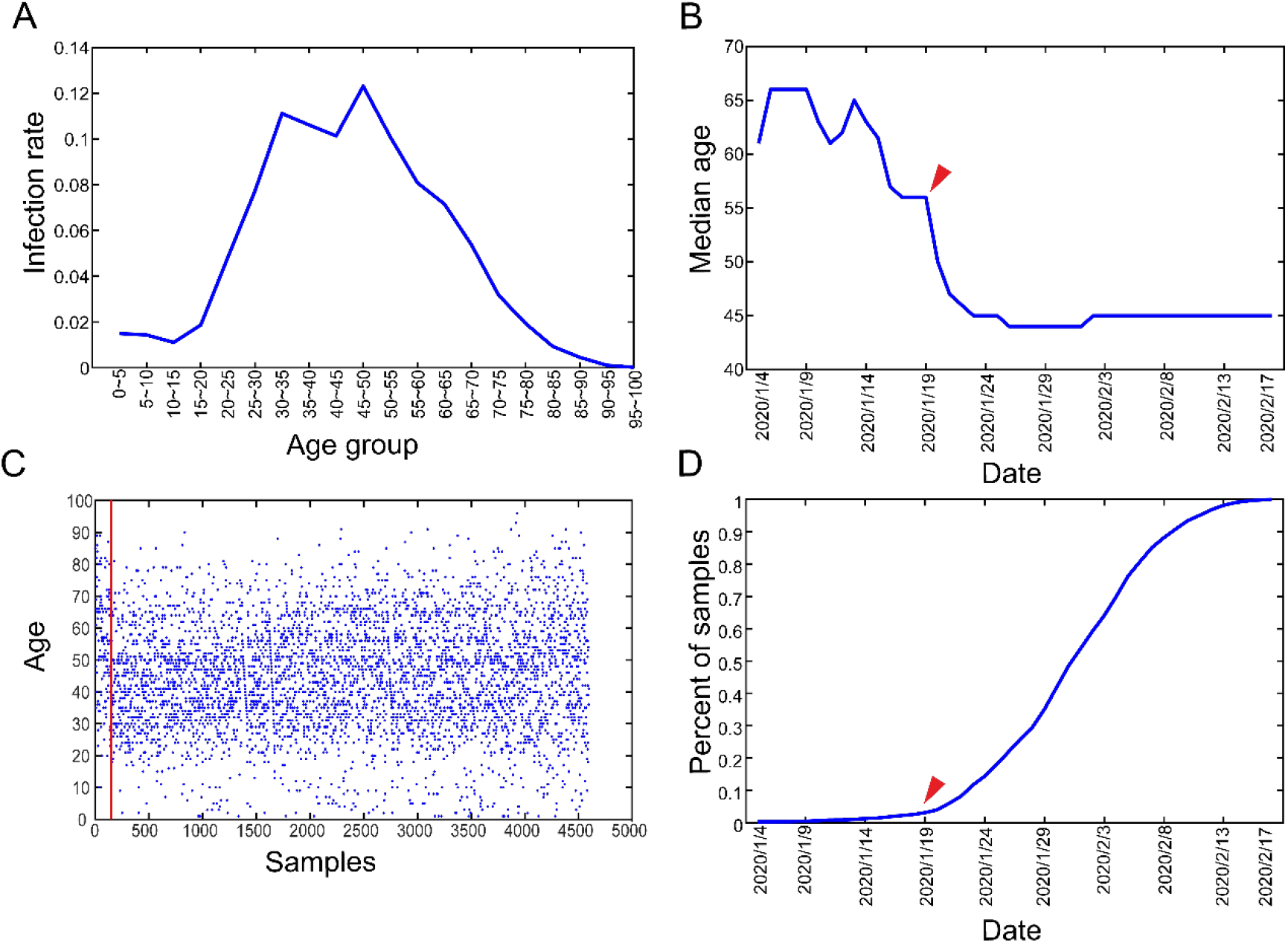
The ages distribution of the COVID-2019 infected samples reported from 26^th^ Nov 2019 to 17^th^ Feb 2020. In figure 1A, the 0∼5 group contains the samples whose ages are larger than zero and smaller than or equal with five. The other group groups samples in the same way; in figure 1C, the red line separates the samples reported before 19^th^ Jan 2020 (the ones on the left hand of the red line) from the samples reported after 19^th^ Jan 2020 (the ones on the right side of the red line)

### The ages distribution of samples reported before and after 19^th^ Jan 2020 is significantly different

Then we sorted samples according to the dates on which they visited the doctors. For each day, we firstly collected the cases that had visited the doctors until that day, then we calculated the median of the ages of the cases, next we calculated the percent of the cases in all samples. We found the median age of the cases decreases significantly after 19^th^ Jan 2020 (Figure 1B). The ages of samples having visited the doctors before 19^th^ Jan 2020 are significantly higher than the ages of samples visited the doctor after that day (Figure 1C, Wilcoxon rank-sum test, single tail, p<0.001). After 19^th^ Jan 2020, the number of samples was explosively increased (Figure 1D). What happened on 19^th^ Jan 2020? The national health commission of China sent commissioners to each province to help control COVID-2019 infection. On 20^th^ Jan 2020, the Wuhan government broadcasted that 136 people were infected with COVID-2019 during the 18^th^ and 19^th^. Dr. Nanshan Zhong confirmed that COVID-2019 could be transmitted by person-to-person contact in the interview of CCTV. People of China started to realize that an infectious disease is spreading in the country.

### In Hubei and Guangdong, the ages distribution of samples reported before and after 19^th^ Jan 2020 is significantly different

We grouped samples by their residences (provinces). We kept the province-groups with more than 100 samples. We found that, in Zhejiang, Tianjin, Sichuan, Shanghai, Shanxi(陕西), Shandong, Jiangxi, Jiangsu, Hunan, Henan, Hainan, and Guangxi, the ages distribution of samples reported before and after 19^th^ Jan 2020 shows no difference (Figure 2, Wilcoxon rank-sum test, single tail, p> 0.05). The ages of samples reported before 19^th^ Jan 2020 are significantly higher than the ages of samples reported after that day in Hubei and Guangdong (Figure 3, Wilcoxon rank-sum test, single tail, Hubei: p<0.001, Guangdong: p<0.0018). Since we have known that older people have a lower COVID-2019 infection rate, more older samples were reported before the 19^th^ Jan 2020 suggests that many middle-aged samples were unfound before that day. The COVID-2019 might be spreading quietly in Hubei and Guangdong before 19^th^ Jan 2020.

**Figure 2.**
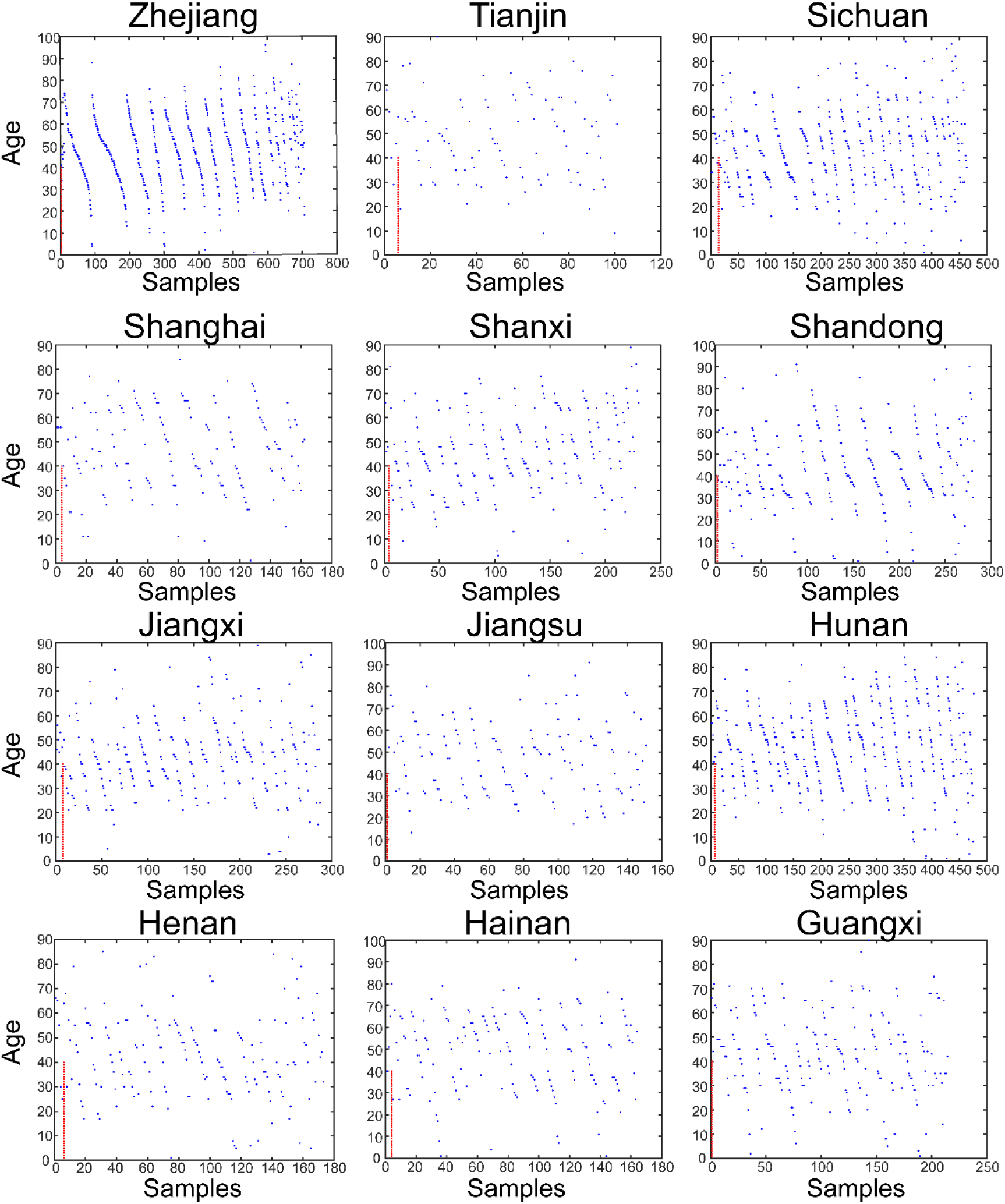
The ages distribution of the COVID-2019 infected samples reported before and after 19^th^ Jan 2020 in the provinces of Zhejiang, Tianjin, Sichuan, Shanghai, Shanxi(陕西), Shandong, Jiangxi, Jiangsu, Hunan, Henan, Hainan, and Guangxi. The red line separates the samples reported before 19^th^ Jan 2020 (the ones on the left hand of the red line) from the samples reported after 19^th^ Jan 2020 (the ones on the right side of the red line)

**Figure 3.**
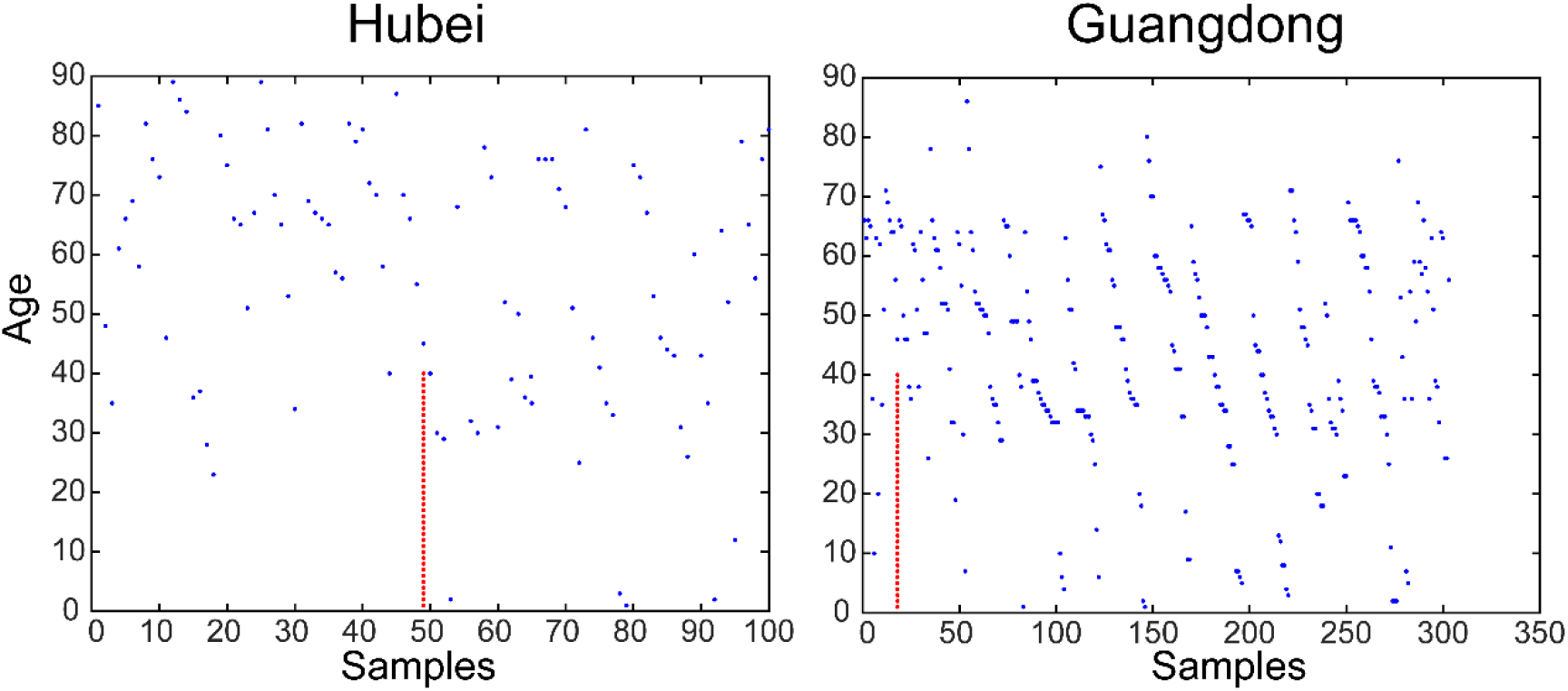
The ages distribution of the COVID-2019 infected samples reported before and after 19^th^ Jan 2020 in the provinces of Hubei and Guangdong. The red line separates the samples reported before 19^th^ Jan 2020 (the ones on the left hand of the red line) from the samples reported after 19^th^ Jan 2020 (the ones on the right side of the red line)

### The ages distribution of each-day-reported samples might serve as a warning indicator of whether all potential COVID-2019 infected cases are found

We took 19^th^ Jan 2020 as the burst day. We collected the samples that had visited the doctors before 19^th^ Jan 2020. Next, we plotted the boxplot of their ages (Figure 4). We selected the days after 19^th^ Jan 2020. Then, for each day, we collected the samples that had visited the doctors on that day and plotted the boxplot of their ages (Figure 4). We found that the ages of samples reported before 19^th^ Jan 2020 tend to be higher than the ages of each-day-reported samples, excluding the ages of samples reported on 14^th^ Feb 2020 (Figure 4, Wilcoxon rank-sum test, single tail, p< 0.05). We infer that when the spreading of COVID-2019 is confirmed in a region and a significant number of COVID-2019 infected cases are reported each day, the ages of each-day-reported cases should be lower than the ages of COVID-2019 infected cases reported before the burst day. The ages of each-day-reported COVID-2019 infected cases have the potency to serve as a warning indicator of whether all potential COVID-2019 infected cases are found.

**Figure 4.**
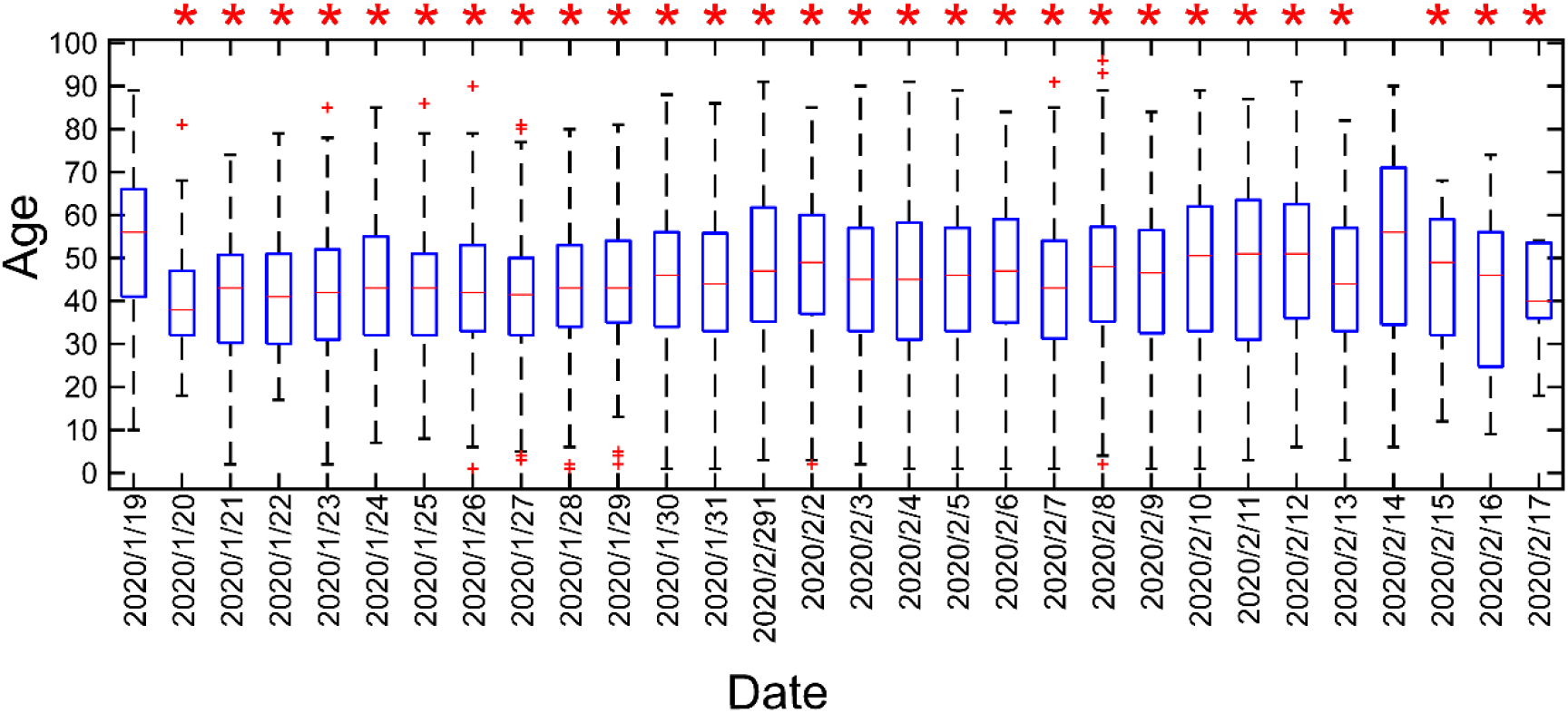
The ages distribution of the each-day-reported COVID-2019 infected samples in China. The group of “2020/1/19” collects the samples reported from 26^th^ Nov 2019 to 19^th^ Jan 2020. The other group collects the samples on the day. “*” indicates that the ages of the samples on the day are significantly lower than the ages of the samples of the “2020/1/19” group.

## Discussion

Based on 1. the fact that the older people have lower COVID-2019 infection rates compared to middle-aged persons and 2. the common sense that the older people prefer to report their illness and get treatment from the hospital, we propose a hypothesis that when the spreading of COVID-2019 is overlooked, more older cases will be reported. We demonstrated the hypothesis with 4597 COVID-2019 infected samples reported from 26^th^ Nov 2019 to 17^th^ Feb 2020 across the mainland of China. The comparison of the ages of the samples reported before and after 19^th^ Jan 2020 shows that COVID-2019 might not spread in most provinces of China before 19^th^ Jan 2020, but it might have spread in Hubei and Guangdong before that day.

This empirical analysis is data-driven. The power of the analysis is limit because only ∼5% of the COVID-2019 infected people in China are included in the study. However, the ages of each-day-reported COVID-2019 infected cases have the potency to serve as a warning indicator of whether all potential COVID-2019 infected cases are found. We believe our work shows some value for its ability to estimate the unfound COVID-2019 infected people.

## Data Availability

All the data can be freely downloaded from the URL provided.

## Contributorship statement

CW designed and carried out the study. MZ improved the study. CW and MZ drafted the manuscript. All authors read and approved the final manuscript.

## Acknowledgments

This article is dedicated to the lovely people who are fighting against COVID-2019

